# Prospective evaluation of genome sequencing versus standard-of-care as a first molecular diagnostic test

**DOI:** 10.1101/2020.09.03.20181073

**Authors:** Deanna G. Brockman, Christina A. Austin-Tse, Renée C. Pelletier, Caroline Harley, Candace Patterson, Holly Head, Courtney Elizabeth Leonard, Kimberly O’Brien, Lisa M. Mahanta, Matthew S. Lebo, Christine Y. Lu, Pradeep Natarajan, Amit V. Khera, Krishna G. Aragam, Sekar Kathiresan, Heidi L. Rehm, Miriam Udler

## Abstract

**Purpose:** To evaluate the diagnostic yield and clinical utility of clinical genome sequencing (cWGS) as a first genetic test for patients with suspected monogenic disorders.

**Methods:** We conducted a prospective randomized study with pediatric and adult patients recruited from genetics clinics at Massachusetts General Hospital who were undergoing planned genetic testing. Participants were randomized into two groups: standard-of-care genetic testing (SOC) only or SOC and cWGS.

**Results:** 204 participants were enrolled and 99 received cWGS. cWGS returned 23 molecular diagnoses in 20 individuals: A diagnostic yield of 20% (20/99, 95%CI 12.3-28.1%)), which was not significantly different from SOC (17%, 95%CI 9.7%-24.6%, P=0.584). 19/23 cWGS diagnoses provided an explanation for clinical features or were considered worthy of additional workup by referring providers. While cWGS detected all variants reported by SOC, SOC failed to capture 9/23 cWGS diagnoses; primarily due to genes not included in SOC tests. Turnaround time was significantly shorter for SOC compared to cWGS (33.9 days vs 87.2 days, P<0.05).

**Conclusions:** cWGS is technically suitable as a first genetic test and identified clinically relevant variants not captured by SOC. However, further studies addressing other variant types and implementation challenges are needed to support feasibility of its broad-scale adoption.

## Introduction

Currently, diagnostic standard-of-care (SOC) genetic testing practices are guided by specialty-based practice guidelines and clinical judgement^1–3^. These practices may consist of a combination of methods such as karyotyping, array-based comparative genomic hybridization, single gene analysis, and multi-gene panels^4^. While high-coverage targeted sequencing technology has broadened the ability to assess and interpret the human genome, this approach has three key limitations. First, it requires that a set of genes be prespecified for each disease area; second, it limits the ability to reanalyze the data after new gene-disease associations are made; and third, it requires provider awareness and commercial availability of numerous disease-specific testing options.

In contrast to disease-focused genetic analysis, exome (WES) and genome sequencing (WGS) have the potential to overcome the limitations of SOC and serve as effective diagnostic tools for rare genetic disorders in children^5–10^. Furthermore, WGS provides more uniform coverage of the genome, expands the scope of variants that can be identified based on documented medical and family history, and can reduce the number of genetic tests necessary to reach a diagnosis^11^.

In this prospective randomized study, we aimed to 1) assess diagnostic yield and clinical utility of clinical genome sequencing (cWGS) across various disease phenotypes and ages at diagnostic evaluation, and 2) explore the challenges associated with implementing cWGS as a diagnostic tool for patients with suspected genetic conditions. Here we report on diagnostic yield, clinical utility, and turnaround time (TAT) of cWGS as compared to SOC.

## Methods

### Study design and participants

Participants were recruited at the time of their clinical genetics evaluation at Massachusetts General Hospital (MGH) in Boston, Massachusetts between March 2018 and July 2019. Six MGH clinics participated in this study: the Cardiovascular Genetics Program (CGP), Medical Genetics Program (MGP), Ataxia Genetics Unit-Neurology (ATX), Gastrointestinal Cancer Program (GIC), Endocrine Genetics (END), and Pulmonary Genetics Clinic (PUL).

To be eligible for the study, patients were required to be pursuing a *diagnostic* genetic test at the time of enrollment; individuals were not eligible if they previously pursued genetic testing for the same indication. Potential participants were identified through medical record review by a research study coordinator and eligibility was confirmed by a study genetic counselor and the referring clinician. Given prior data on the utility of sequencing pediatric patients and their parents^12^, patients under the age of 18 were offered enrollment as a family trio. Eligibility criteria are further described in Table S1. Consent sessions with a genetic counselor involved a discussion of study logistics, an overview of cWGS, and potential results, which included both primary and non-primary findings. Participants were allowed to opt-out of receiving results in the American College of Medical Genetics minimum list of genes to be reported as secondary findings (ACMG-59)^13^. After enrollment, patient features were abstracted from electronic medical records (EMR) and recorded as Human Phenotype Ontology (HPO) terms using PhenoTips^14^.

Randomization was used as a strategy to avoid influencing the referring provider’s SOC approach and biasing patient choices for reflex testing. Enrolled participants were randomized 1:1 to receive only SOC or to receive both SOC and cWGS. Referring clinical providers, study staff members with patient interaction, and patients were blinded to randomization status until cWGS report availability of three months after enrollment if randomized to the control arm. Block randomization stratified by clinic was implemented to ensure that a comparable proportion of individuals from each clinic received cWGS. Participants enrolled as a family trio were randomized independent of the clinic in which they were enrolled.

All participants were asked to complete two surveys – one at the time of enrollment and one after learning their randomization status and receiving cWGS results if enrolled into the cWGS arm. This study was completed as a demonstration project in the MGH Center for Genomic Medicine and was approved by the Mass General Brigham Institutional Review Board.

### Genome Sequencing, Analysis, and Reporting

Genome sequencing was performed in the Clinical Laboratory Improvement Amendments-certified, CAP-accredited Clinical Research Sequencing Platform at the Broad Institute of Massachusetts Institute of Technology and Harvard in Cambridge, MA (CLIA#22D2055652). All samples achieved a minimum coverage of 20 reads per base for at least 95% of the genome, with a minimum mean coverage across the genome of 30 reads per base.

The Partners Laboratory for Molecular Medicine (CLIA#22D1005307) performed sequence realignment, variant calling, annotation, and report generation. Reads were aligned to the human reference sequence (GRCh37) using Burrows-Wheeler Aligner (BWA), and variant calls were made using the Genome Analysis ToolKit (GATK). cWGS analysis methods and reporting criteria are described in the Supplementary Methods and Figure S1.

The cWGS data was retrospectively screened for SOC-reported copy number variants as described in the Supplementary Methods.

### Molecular Diagnosis and Clinical Utility

In this study, sequencing results were categorized as a ‘molecular diagnosis’ if they met all of the following criteria: (1) variant(s) classified as Pathogenic, ‘P’ or Likely Pathogenic, ‘LP’, (2) variant(s) in genes with known disease association, and (3) variant(s) in allele states consistent with the inheritance pattern of the associated disorder. Further, molecular diagnostic findings were categorized as ‘primary’ if they explained or partially explained the indication for SOC. ‘Non-primary’ molecular diagnoses were those that were unrelated to the patient’s indication for testing but were related to the patient’s family history, explained an additional phenotype identified upon EMR review, or were indicated on the ACMG-59 secondary findings list.

The molecular diagnostic yield of SOC was compared to that of cWGS for all patients who received both SOC and cWGS reports. All molecular diagnoses on cWGS were evaluated for ‘clinical utility’. To assess clinical utility, we evaluated if the result provided a diagnosis consistent with the patient’s reported phenotype and if the result informed medical management; clinical utility was confirmed by the referring clinician.

Turnaround time was determined by calculating the number of days from the test order date to the date the report was generated. When multiple tests were performed as part of the SOC process (i.e. microarray plus WES), turnaround time was designated as the number of days between the first test order date to the date of last report.

### Statistical analyses

Mean values between groups were compared using the two-sample t-test. Comparison of multiple values between the two study arms was performed using two-way analysis of variance (ANOVA). Diagnostic yields were compared using the two-sample test of proportions. Statistical significance threshold was set at alpha = 0.05. All analyses were performed in Stata/IC 14.2.

## Results

### Participant demographics, clinics of enrollment, and genetic test indications

Between March 2018 and July 2019, 3,771 patients were evaluated by one of the six participating MGH genetics clinics; 204 patients were enrolled and 100 were randomized to receive cWGS (Fig. 1, Figure S2). One participant did not receive SOC due to insurance challenges and was removed from subsequent analysis -- this resulted in 99 participants who received both SOC and cWGS. The highest volume enrollment sites were the Cardiovascular Genetics Program (n=69, 34%) and Medical Genetics Program (n=60, 29%) (Table 1).

**Fig. 1.**
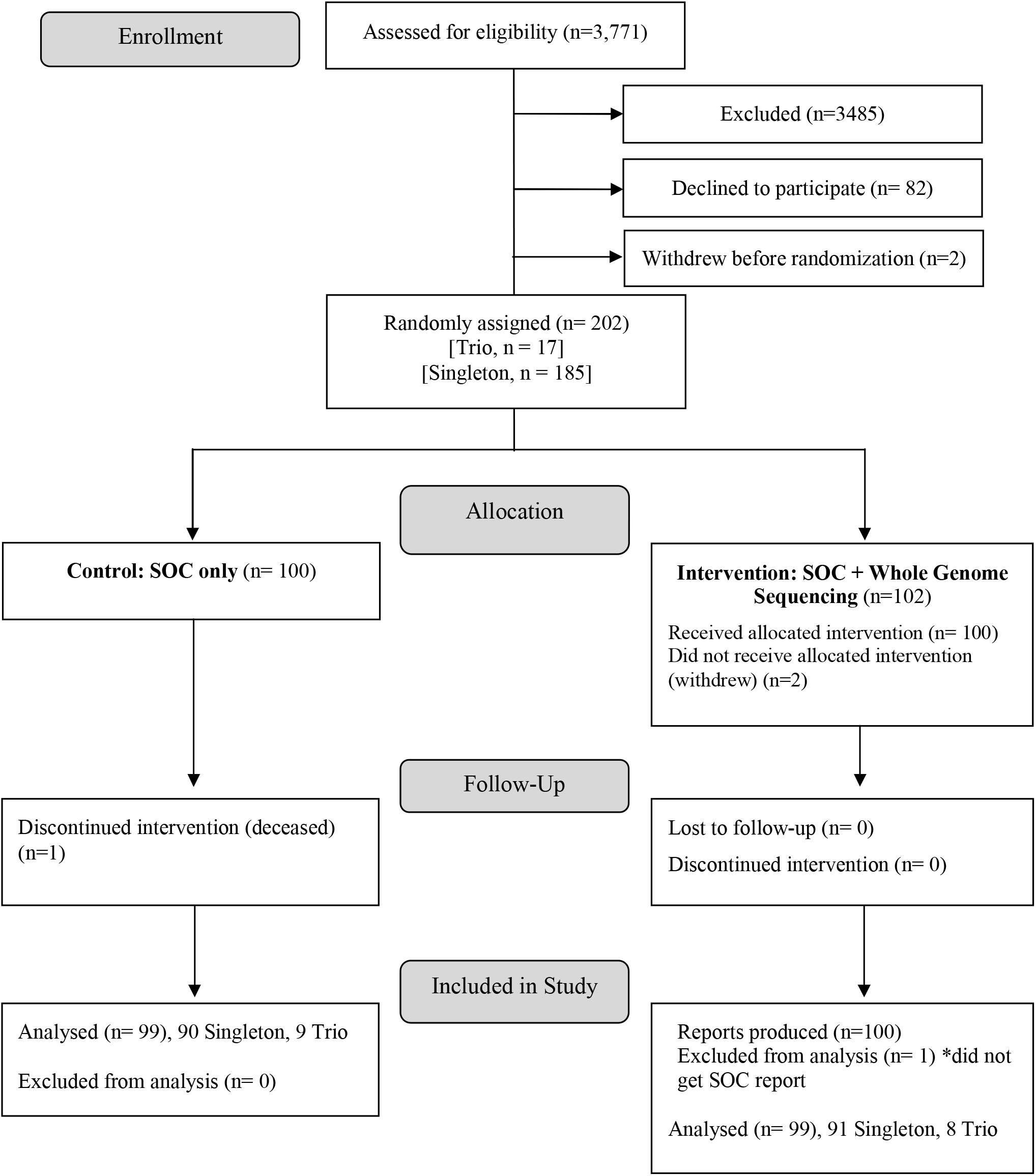
Proband participant enrollment flowchart. Note: 2 additional cWGS reports were produced for parents in a trio, but were not included in this diagram.

**Table 1.**
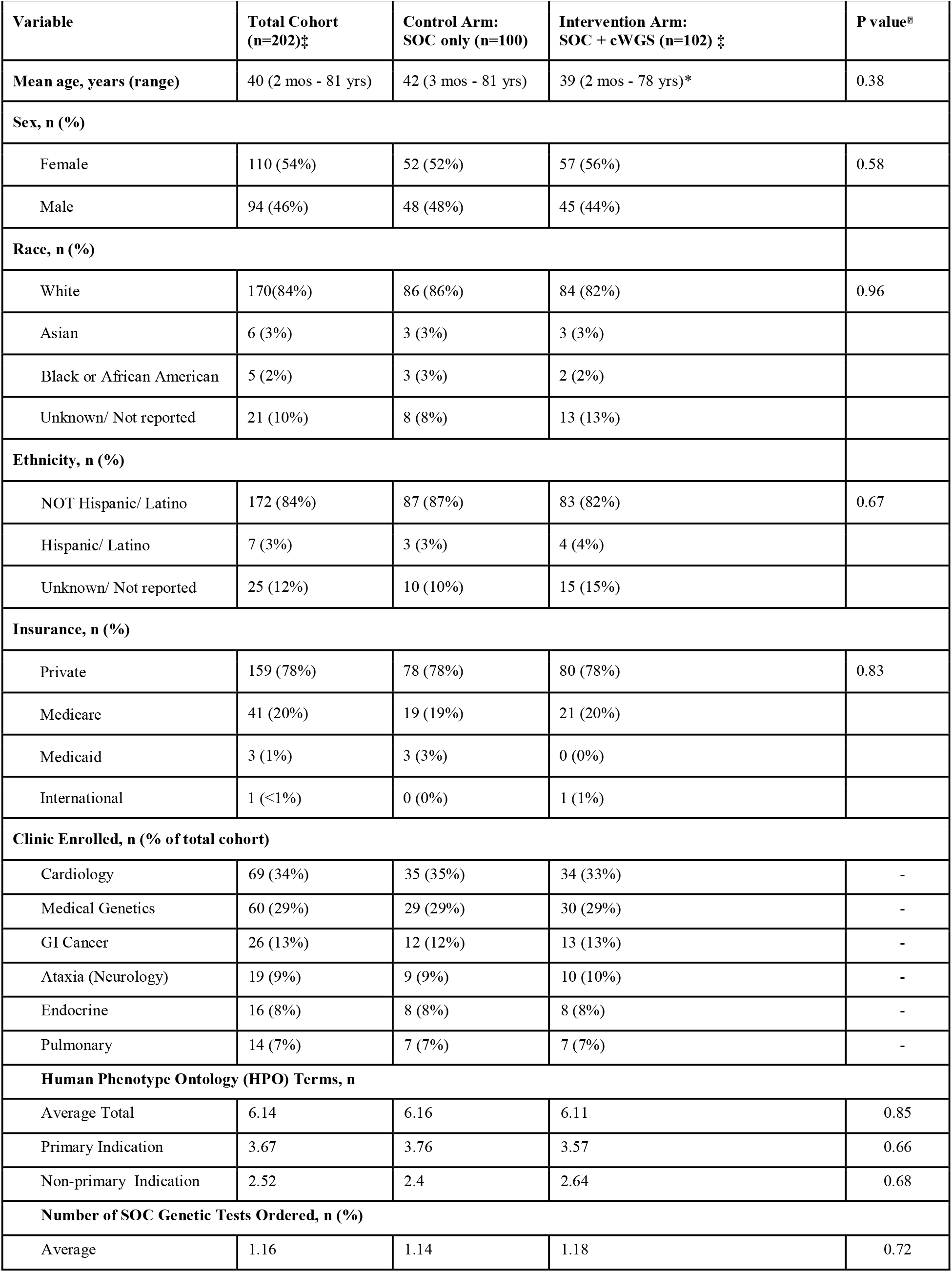
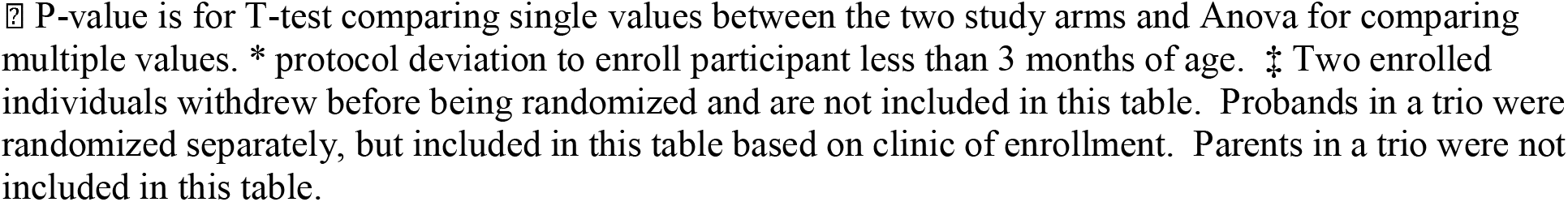
Participant Characteristics and Enrollment.

The average age of the total cohort was 40.1 years, with 82% (n=168) age 18 years or older. The majority of all participants (82%) were White (Table 1). 17/36 pediatric probands were enrolled as a trio with both biological parents. The most common SOC test ordered was a multi-gene panel (n = 137, 65%) (Table 1, Figure S3). The average number of HPO terms per participant was 6.14 (Table S2). No statistically significant differences in age, sex, race, ethnicity, insurance, HPO terms, or number of SOC tests ordered were observed between the control (SOC only) and intervention (SOC + cWGS) groups (P-values > 0.05, Table 1).

### Molecular diagnostic yield: Genome Sequencing (cWGS)

The overall cWGS molecular diagnostic yield was 20% (20/99 participants who received both SOC and cWGS, 95%CI 12.3-28.1%). Given the expanded analytical scope of cWGS, some individuals received multiple diagnoses, yielding a total of 23 molecular diagnoses. The molecular diagnostic yield for pediatric participants (<18 years of age) was 26.3% (5/19) and there was no significant difference in yield between singleton and family trio cWGS (singleton: 27.3% (3/11), trio: 25% (2/8), P value > 0.05). The cWGS molecular diagnostic yield for adult participants was 18.9% (15/80), ranging from 0% (0/12) in the Gastrointestinal Cancer Clinic to 30% (3/10) in the Ataxia Unit-Neurology (Fig. 2).

**Fig. 2.**
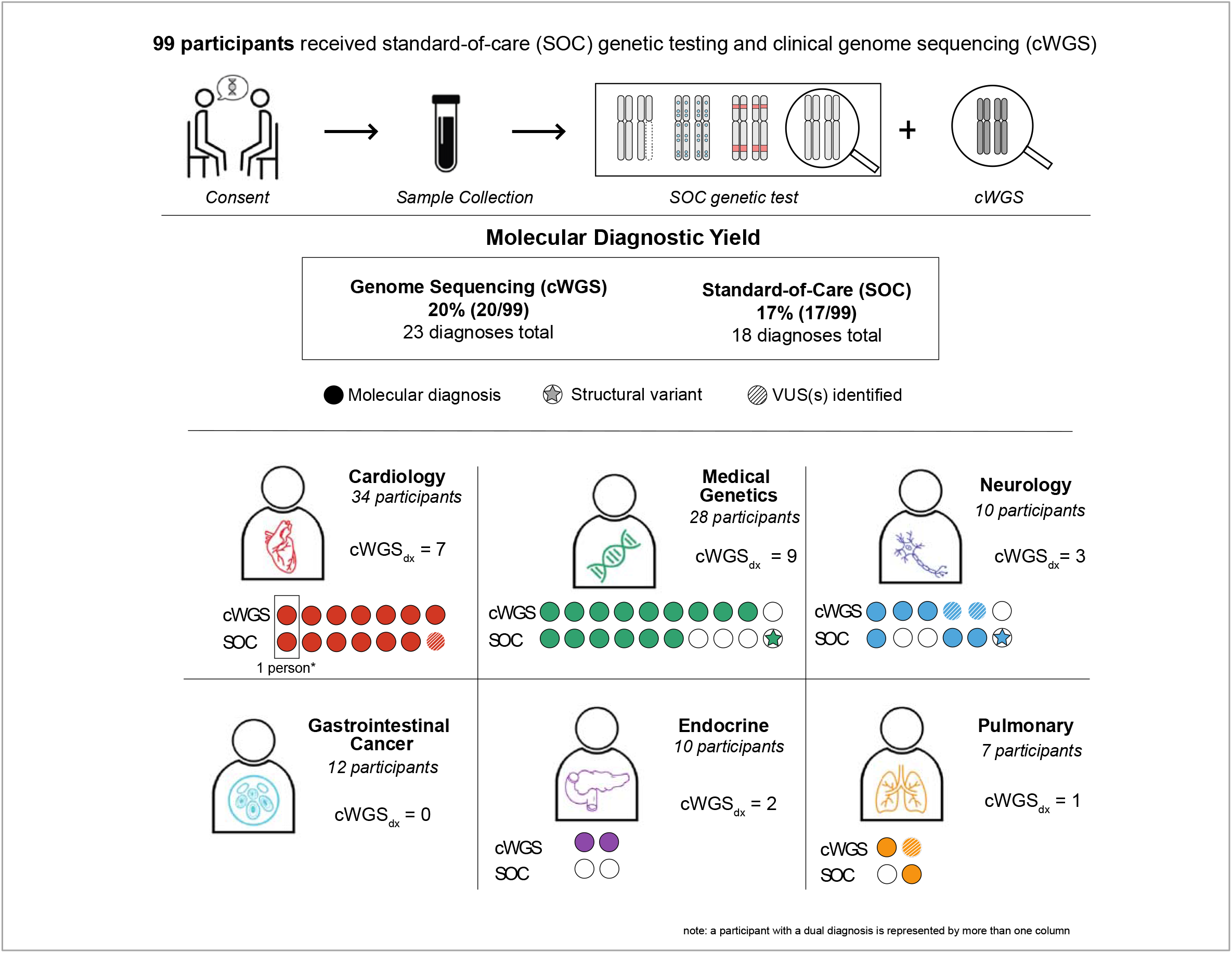
Molecular diagnoses (probands only) made by cWGS and SOC. *A participant with multiple diagnoses is represented by more than one column.

### Comparison of variants detected on SOC vs. cWGS

All 23 of the P/LP small (<20bp) sequence variants reported by SOC were technically detected by cWGS and filtered appropriately, corresponding to a sensitivity of 100%. (Table S3). In addition to sequence variants, 3 P/LP copy number variants (CNVs) were reported by SOC (Table S3, Cases 204CGS, 152CGS, 170CGS). Although CNV calling had not yet been validated in the cWGS test used for this study, retrospective analysis using a research-grade WGS CNV calling algorithm detected all 3 CNVs identified by SOC (Table S4).

### Molecular diagnostic yield: cWGS vs. SOC

SOC reported 56.5% (13/23) of the molecular diagnoses reported on cWGS. Due to differences in variant classification, one cWGS molecular diagnosis of *MYH7*-related hypertrophic cardiomyopathy was detected but not categorized as a molecular diagnosis on SOC (Patient 35CGS, Tables S5 and S6). As a result, when disregarding classification discrepancies, SOC reported 60.9% (14/23) of the cWGS diagnoses. SOC delivered a total of 18 molecular diagnoses in 17 individuals (Fig. 2)-- a molecular diagnostic yield of 17% (17/99 participants, 95%CI 9.7%-24.6), which was not significantly different from cWGS (P=0.584). Similar to cWGS, SOC diagnostic yield was lowest in the gastrointestinal cancer (0%; 0/12 participants) and endocrine (0%; 0/10 participants) clinics, and highest in the Ataxia Unit-Neurology (30%; 3/10 participants).

Amongst the participants who received both SOC and cWGS, nine molecular diagnoses were detected only by cWGS (Fig. 2, Table S5, S6): four diagnoses met study criteria for primary diagnosis and five met study criteria for non-primary phenotype diagnosis. For two of the cWGS primary molecular diagnoses, reported variants were in genes not included in the SOC analysis (Patients 32CGS, 65CGS – see case vignette below; Table S5, S6). The other two cWGS primary molecular diagnoses were attributed to variants that were detectable by the SOC method (WES) but were not reported on SOC due to differences in laboratory reporting practices (Table S5, S6; Patients 63CGS, 80CGS).

> ***Case 65CGS:** A child presented to the Medical Genetics Program for evaluation due to delayed speech and language development, and autistic behavior. At the time of her visit, three tests were ordered -- Fragile X, Autism/ID Panel, and Microarray -- all were negative. This patient was enrolled in the study as a family trio. cWGS revealed two pathogenic* ***GJB2*** *variants (p.Gly12ValfsX2 andp.Ser139Asn) confirmed in trans, suggesting a diagnosis of autosomal recessive deafness. Upon review of this result with the family, it was uncovered that the patient never had a hearing evaluation (including a newborn hearing screen). This case highlights the potential of genome sequencing to (1) reduce the number of genetic tests ordered and (2) make a diagnosis based on expanded phenotyping and analysis*.

The remaining five molecular diagnoses captured exclusively by cWGS included four conditions with clinical features that had possible relevance to the probands’ non-primary phenotypes (Patients 159CGS, 169CGS, 29CGS, 32CGS, Table S5, S6), and one that was relevant to family history only (patient 170CGS, Table S5, S6). Non-primary phenotypes and family history were not the focus of the SOC approaches. As a result, these genes were not included in SOC ordered.

It should be noted that five molecular diagnoses were made by SOC but not cWGS (Fig. 2). Three of the diagnoses made only by SOC were the result of variant classification differences (63CGS- *TOR1A, 80CGS-NPC1*, 196CGS- *PIK3CA*, Table S5, S6). For the remaining two cases (Cases 204CGS, 152CGS), the molecular diagnostic discrepancy can be attributed to the identification of a CNV by SOC, which were only retrospectively examined in this first pilot of cWGS.

cWGS and SOC reports also differed in reporting of non-diagnostic variants. Amongst 99 participants who received both SOC and cWGS, 57 VUS were reported on SOC and/or cWGS (Table S6). Five VUSs identified exclusively by cWGS in five participants prompted additional clinical workup (Fig. 3). Two case examples are described below -- in both cases, familial testing was recommended to determine the phase of the identified variants; this testing was still pending at the time of this publication.

> ***Case 9CGS:** Two VUSs,c.3855C>T (p.Ile1285Ile) and c.2097+3_2097+15del (p.?), in the* ***SYNE1*** *gene were detected by cWGS in a proband who was referred for SOC triplet repeat expansion testing based on his presentation of cerebellar atrophy, diplopia, and mild speech impairment. These phenotypes are consistent with a diagnosis of autosomal cerebellar ataxia type 1, which is caused by pathogenic variation in the SYNE1 gene. Both identified variants were classified as variants of uncertain significance due to limited available data. Given the close match in phenotype, and presence of two extremely rare variants, suspicion was higher for diagnostic relevance*.
>
> ***Case 163CGS:** In a proband with ataxia, abnormal MRI, dysarthria, and a personal and family history of basal cell carcinoma, cWGS identified one pathogenic variant(p.Arg616Pro) and one variant of uncertain significance (p.Gly413Val) in the ERCC2 gene, which is associated with a spectrum of autosomal recessive conditions including trichothiodystrophy, xeroderma pigmentosum, and Cockayne syndrome. Notably, at least 25% of individuals with ERCC2-related disorders have progressive neurologic abnormalities, including ataxia and neurodegeneration in the cerebrum and cerebellum*.

**Fig. 3.**
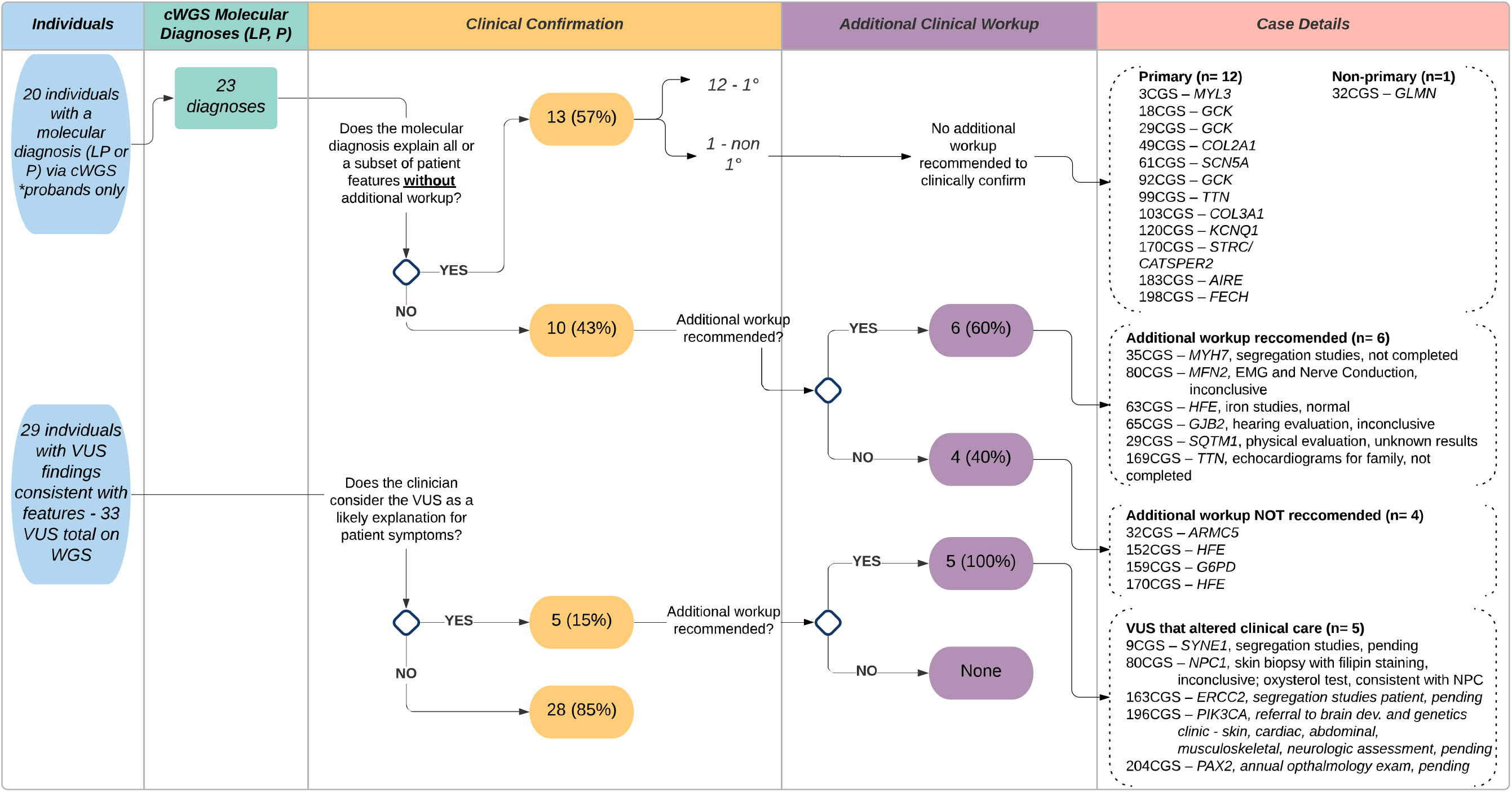
Clinical utility of cWGS molecular diagnoses and suspicious VUS results.

**Fig. 4.**
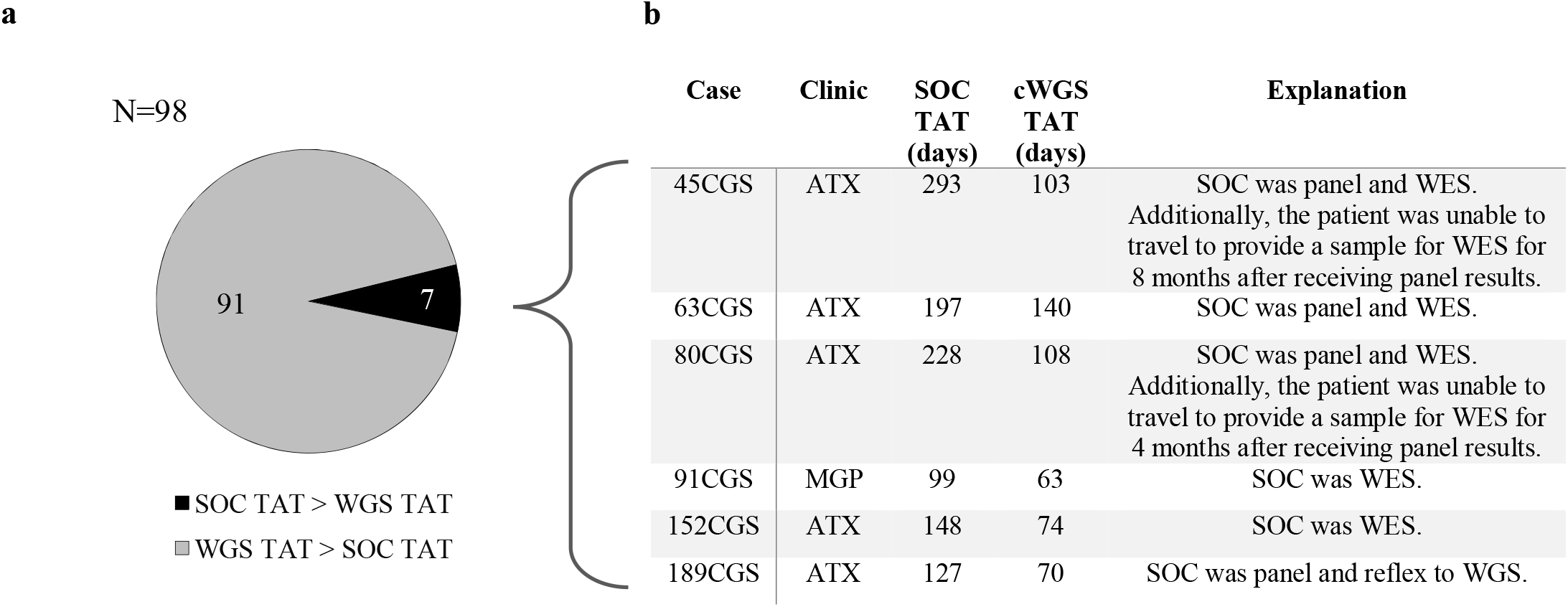
Comparison of turnaround time: SOC vs. cWGS. (a) Number of cases where turnaround time (TAT) for a SOC report was longer than that of WGS and vice versa. (b) Specific examples when a cWGS report was available before the SOC diagnostic pipeline was complete. Note: One case was removed from this analysis because the report date was unknown. In another case, turnaround time (TAT) was adjusted due to insurance concerns resulting in a delay in the start of the SOC genetic testing pipeline.

### Clinical Utility & Impact on management

Upon review of post-clinic notes and/or discussions with referring providers, 13 of 23 (57%) cWGS molecular diagnoses explained current clinical features or a subset of features without additional workup – 12 were related to the primary indication for testing and one was related to a non-primary indication (Fig. 3). Of the remaining 10 cWGS molecular diagnoses with unclear clinical utility, referring providers recommended additional workup for six cases, including electromyography (EMG), hearing evaluation, and iron studies. Molecular diagnoses have not yet been clinically confirmed based on additional workup for these cases.

To further explore the medical importance of cWGS results, we reviewed the utility of clinically suspicious VUS findings. Despite uncertain variant pathogenicity, referring clinicians reported that they planned to change medical management and/or pursue additional workup for five patients with VUSs based on cWGS results (Fig. 3). To date, a diagnosis of Niemann Pick Type C was confirmed based on additional workup for one patient (Case 80CGS).

> ***Case 80CGS:** A female in her 40s presented to the Ataxia Unit for evaluation due to ataxia, cerebellar atrophy, dysphagia, and dysarthria. At the time of her visit, an autosomal dominant triplet repeat ataxia panel was ordered and was negative. WES was pursued by the clinical team in follow up to these results, which identified two variants (p. Gln438X, P andp.Phe68del, LP) in* ***NPC1****, suggesting a diagnosis of Niemann-Pick Disease Type C. In parallel, genome sequencing identified the same variants in NPC1, however the variant classification differed (LP and VUS). Additionally, genome sequencing revealed a* ***MFN2*** *variant(p. Arg707Trp, LP), suggesting a diagnosis of Charcot-Marie Tooth Type 2A. Follow-up with the WES testing lab revealed that the MFN2 variant was detected but not reported due to a perceived lack of relevance to the patient phenotype. Upon review with the referring clinical team, additional workup was recommended, including: (1) skin biopsy with filipin staining to evaluate for Niemann-Pick Disease Type C -- inconclusive (approximately 50% staining) and (2) electromyography (EMG) and nerve conduction studies (NCS) to evaluate for Charcot-Marie Tooth Type 2A, which were inconclusive. Subsequently, the patient received an oxysterol test, which was consistent with a diagnosis of Niemann-Pick Disease Type C. She is now taking Miglustat to stabilize and slow progression of the disease. This case highlights the (1) inconsistencies in exome/genome reporting standards, and (2) need for clinical correlation of molecular diagnoses made on exome and genome sequencing*.

cWGS also confirmed one clinical diagnosis of hemochromatosis in a parent enrolled in this study as a part of a family trio. In total, 15 cWGS molecular diagnoses were confirmed by clinical workup; two (170CGS parent, 32CGS) would not have been made by standard-of-care genetic test approaches.

### Diagnostic workup timeline

On average, 1.16 genetic tests per patient were ordered during SOC workup (Fig. 1, Table 1). No differences were observed in number of SOC genetic tests ordered between study arms (P-value > 0.05), suggesting that enrollment in the study did not impact the SOC approach. Amongst participants who received cWGS (n=99), TAT data was available for 98 participants for at least 11 months post-enrollment. The average time from the first SOC test order date to last available SOC report was 33.9 days (minimum = 7, maximum = 293, 95% CI, 24.5-43.3) and 87.2 days (minimum = 49, maximum = 162, 95% CI, 82.5-91.9) via cWGS. The SOC workup timeline exceeded cWGS for 6 cases, all of which involved exome or genome sequencing – 5 of these 6 were recruited from the Ataxia clinic. Notably for the case with the longest SOC TAT (293 days), the patient was unable to return to care for 8 months between tests, exaggerating the TAT. When comparison was restricted to cases that reached a diagnosis, TAT for cWGS was still significantly longer than for SOC (SOC, 51.2 days, cWGS 92.3 days, P-value <0.05). However, it should be noted that the cWGS TAT may not be reflective of commercially available WGS tests given the limited WGS analysis staff available for this study.

## Discussion

Previous studies suggest that exome/genome sequencing be utilized as the first genetic test for individuals with a suspected genetic disorder, citing increased diagnostic yield, reduced time to reach a diagnosis, and economic advantages over the SOC step-wise approach to genetic testing^15,16^. These studies predominantly enrolled pediatric patients or focused on a specific disease area. To our knowledge, this is the first prospective study comparing the diagnostic yield and clinical utility of singleton and family-trio clinical genome sequencing to that of SOC practices across age groups and medical specialties. This study was particularly unique in that genome analysis and interpretation was conducted within an integrated healthcare setting, allowing for collegial discussions about the significance of cWGS results with referring providers.

cWGS resulted in a molecular diagnostic yield of 20% (26.3% in pediatrics and 18.9% in adults); this yield was consistent with other studies that report diagnostic yields ranging from 14% – 76%^15,17,18^. Of note, the clinic with the highest cWGS diagnostic yield in this study was the adult ataxia unit (neurology) at 30% (3/10) – all diagnoses were due to the identification of sequence variants. This was an unanticipated finding as 74% of the SOC genetic tests were ordered on the basis of concern for a triplet repeat expansion disorder (Figure S3). Upon further review of previous studies, the use of WES/WGS has been suggested as a way to improve diagnostic yield for adults with clinically heterogeneous cerebellar ataxias, with yields ranging from 21-46%^19-22^. Despite equivalent SOC and cWGS diagnostic yields observed in this clinic, cWGS identified clinically suspicious VUS results not assessed by SOC in two additional participants from the ataxia unit (cases 9CGS, 163CGS), further supporting this recommendation.

The TAT for SOC was shorter than cWGS for 93.9% (92/98) of cases. However, this largely reflected the limited staff dedicated to case analysis. Recently, optimized sample preparation, sequencing, and data processing steps and artificial intelligence-assisted analyses have been reported to reduce cWGS TAT to less than 30 hours^23^. If cWGS is to be implemented as a first-line test, it is feasible that investment in infrastructure to support rapid analyses could make TAT comparable to or significantly quicker than existing SOC testing options.

This study also revealed multiple sources of reporting differences between SOC and cWGS that will require consideration before cWGS is broadly implemented. The identification of diagnostic findings that partially explained participant phenotypes in genes that were omitted from the ordering provider’s SOC workup highlights the advantages of an unbiased approach to genetic testing such as cWGS. However, cWGS also revealed diagnoses that were unrelated to the patient’s primary indication for testing, which may be undesirable for some patients. Another source of reporting differences were owed to discrepancies in variant classification^24^, highlighting the importance of ongoing efforts to refine the ACMG/AMP classification criteria and support data sharing (ClinGen, https://clinicalgenome.org/; ClinVar, https://www.ncbi.nlm.nih.gov/clinvar/). Laboratory reporting practices represented a key third source of discordance between cWGS and SOC reports in this study. It is common practice for targeted sequencing tests to include all variants classified as P, LP, or VUS on the report. Given the drastically increased scope of genomic sequencing, current guidelines for genomic sequencing suggest that VUSs should only be reported in genes highly relevant to the patient phenotype^25^. In following with this practice, several VUSs included on SOC reports were reviewed and excluded from reporting by cWGS due to perceived lack of relevance to the patient phenotype. The observation of fewer reported VUSs together with improved diagnostic yield for cWGS as compared to SOC suggests that more targeted genetic testing reports may be one benefit of widespread implementation of cWGS.

Prospective comprehensive CNV analysis was not performed on the cWGS data in this study. The detection by SOC of clinically significant CNVs suggests that the full potential of cWGS as a diagnostic tool was not realized in this study. However, it is encouraging that 3/3 SOC-reported diagnostic CNVs were detectable in the WGS data using CNV calling algorithms. Future studies will include clinical validation of these algorithms and a comprehensive evaluation of the impact of structural variant calling on the diagnostic yield of cWGS.

We would be remiss not to note that this study was limited by multiple systemic barriers that impact access to and uptake of genetic services and testing within a healthcare system. In a 2015 systematic review these obstacles to genetic services were identified, which included: lack of awareness of personal/ patient risk factors, lack of knowledge of family medical history/ lack of obtaining adequate family history, and lack of knowledge of genetic services^26^. These factors influenced the patients identified and recruited for this study and negatively impacted participant diversity. Beyond access to genetic services, uptake of SOC appointments and testing was a barrier to participation. To participate in this study, individuals were required to attend an in-person appointment and pursue SOC at the time of enrollment. Given that 189 eligible patients did not attend their SOC appointment and a portion of eligible patients deferred SOC genetic testing due to insurance coverage concerns, patients were likely excluded from the study due to challenges preventing them from traveling to Boston for an appointment as well as underlying insurance challenges imposed by the United States healthcare system (Figure S2). Further, 176 eligible patients were excluded because they were not English speaking, emphasizing the need for dedicated resources to support diverse populations in clinical care and research. Given that cWGS requires a blood sample, we were also limited by our inability to collect parental samples for trio-WGS when both parents were unable to come to clinic – often due to work, travel, and family-related obstacles. In order to equitably offer the most comprehensive cWGS evaluation, effort is needed to develop methods that allow cWGS to be run on saliva or buccal samples which can be submitted remotely. Lastly, this was a hospital sponsored clinical research study. Given that most payers consider cWGS to be investigational, efforts must be made to contract with insurance companies and to conduct the necessary cost-effectiveness analyses needed to improve payer coverage of this test; doing so will make cWGS accessible to more patients.

This study provides evidence that cWGS is suitable as a first-line diagnostic genetic test, regardless of patient age or clinical specialty. However, metrics beyond diagnostic yield and turnaround time need to be considered prior to broad scale implementation. Capturing the full scope of utility and feasibility, with a particular focus on payer coverage, will allow us to move towards equitable and scalable delivery models of genomic medicine.

## Data Availability

De-identified genomic and phenotype data will be made available upon request through a controlled access database. All variants identified by WGS have been submitted to ClinVar by the Partners Healthcare Personalized Medicine Laboratory for Molecular Medicine.

## Acknowledgements

This study was funded by the Department of Medicine at Massachusetts General Hospital. Illumina supplied a portion of the sequencing reagents to enable this study. We thank the patients and families for participating in this study. We are grateful to Stephanie Harris, Lauren O’Grady, Jessica Waxler, Marcie Steeves, Helen Chen, Megan Hawley, Erica Blouch, Linda Rogers, Kristen Shannon, Dr. David Sweetser, Dr. Paula Goldenberg, Dr. Frances High, Dr. Amel Karaa, Dr. Angela Lin, Dr. Inderneel Sahai, Dr. Stephanie Santoro, Dr. Mark Lindsay, Dr. Steven Lubitz, Dr. Christopher-Newton-Cheh, Dr. Jeremy Schmahmann, and Dr. Thomas Bernard Kinane for referring patients to the study. We also thank Dr. Edyta Malolepsza, Dr. Harrison Brand and members of Dr. Michael Talkowski’s laboratory for assisting with SV calling and analysis.

